# *Lactiplantibacillus plantarum* PD01 supplementation reduces microplastics and microplastic-associated chemicals in humans: A randomized, double-blind, placebo-controlled trial

**DOI:** 10.64898/2026.08.12.26360001

**Authors:** Roujun Lu, Long Zhao, Xiaodan Huang, Ruihua Dong

## Abstract

**BACKGROUND:** Plastic products improve daily convenience but release pollutants that threaten human health, and effective elimination strategies remain limited.

**METHODS:** In a randomized, double-blind, placebo-controlled trial, we assessed whether *Lactiplantibacillus plantarum* PD01, previously isolated from a donor with high-microplastic (MP) excretion, accelerates MP elimination. Of 106 participants recruited, 96 were randomized to placebo or PD01. Fecal MPs and urinary phthalate metabolites were measured by pyrolysis-gas chromatography-mass spectrometry and liquid chromatography-mass spectrometry at baseline, week 3, and week 6. Primary outcomes were changes in MP concentrations and phthalate metabolite levels, analyzed with linear mixed-effects models.

**RESULTS:** PD01 significantly increased fecal excretion of polycarbonate (PC) and total MPs at week 3 (PC: estimated mean difference, 3.19; 95% confidence interval [CI], 0.98 to 5.40; interaction P =0.005; total MPs: 1.99; 95% CI, 0.78 to 3.21; interaction P =0.001) and week 6 (PC: 2.87; 95% CI, 0.66 to 5.08; interaction P =0.011; total MPs: 1.25; 95% CI, 0.04 to 2.47; interaction P =0.044). By week 6, urinary phthalate metabolites were significantly lower with PD01 than placebo (MnBP: -0.96; 95% CI, -1.73 to -0.18; interaction P =0.016; MEHHP: -0.91; 95% CI, -1.79 to -0.02; interaction P =0.046; MECPP: -1.06; 95% CI, -2.09 to -0.03; interaction P =0.045; ΣDEHP: -0.97; 95% CI, -1.84 to -0.09; interaction P =0.031).

**CONCLUSIONS:** This trial provides a feasible, low-cost, gut-targeted strategy to accelerate pollutant clearance and represents the first clinical evidence that human plastic burden can be lowered without dietary or behavioral restrictions. (ClinicalTrials.gov: NCT07416214.)

## Introduction

Global plastic production has surged dramatically since 1950s, generating enormous plastic waste that poses multiple crises for ecosystems and human health. Microplastics (MPs), fragments of degraded larger plastics, are now pervasive across natural environments and are inevitably ingested by humans. Extensive evidence has confirmed the presence of MPs in feces, blood, breast milk, and major organs, such as brain, lungs, liver and kidneys ^1^. Toxicological studies indicate that MPs bioaccumulation potentially induces systemic damage through inflammatory responses, oxidative stress, and cellular damage ^2^.

In addition to MPs themselves, chemicals released from MPs, such as phthalate esters (PAEs), are also well-known endocrine disruptors ^3^. Exposure to PAEs elicits multi-organ toxicity, including reproductive disorders, metabolic disruption, liver damage, neurotoxicity, immunotoxicity, and carcinogenicity ^4^. Beyond serving as a source of PAEs release, the hydrophobic surfaces of MPs actively facilitate adsorption of PAEs from the surrounding environment, leading to concurrent exposure to both pollutants ^5,6^. The synergistic toxicity of MPs and PAEs further exacerbates oxidative stress, inflammation, apoptosis, necrosis, and gut microbiota dysbiosis ^7^. The latest cohort research demonstrates that reducing plastic exposure through a low-plastic diet significantly decreases urinary levels of PAE metabolites ^8^. However, strategies to reduce MPs exposure by restricting diet and behavior are cost-prohibitive and impractical, since plastics are now too deeply embedded in daily life to be readily replaced, and the ubiquitous presence of MPs in ecosystems makes exposure unavoidable. Therefore, eliminating MP bioaccumulation from the body is critical for reducing the toxicity of these co-occurring pollutants.

Recently, lactic acid bacteria have been developed as promising agents to mitigate the toxicity of MPs and PAEs. *Lactobacillus* and *Bifidobacterium* species counteract the harmful effects of these pollutants by toxin adsorption and enhanced fecal excretion ^9^. However, these strains were screened from fermented foods and dairy products by evaluating their adsorption capacity *in vitro*; whether they can effectively adsorb and remove pollutants in the human body remains unknow. Furthermore, these preclinical studies are typically conducted using acute, high-dose rodent experiments, which far exceed realistic human exposure levels at concentration, pollutant diversity, and exposure patterns ^10,11^. Consequently, empirical evidence is still lacking regarding whether these strains, once colonized in the human gut, can effectively mitigate toxicity *in vivo*. Human clinical trials are therefore necessary to validate the efficacy of candidate strains for real-world application.

To address these limitations, we previously isolated *L. plantarum* PD01 from individuals with high MP excretion capacity, and validated its ability to alleviate MP-induced toxicity in a murine dietary mixture exposure model ^12^. The *L. plantarum* PD01 has been found to prevent MPs translocation into tissues, reduce residues of PVC, PET, PS and PE in the intestinal tract of exposed mice, and alleviate MP-induced inflammation, obesity and gut microbiota dysbiosis. To further evaluate its effects in human, we designed the first randomized, double-blind, placebo-controlled trial to determine the efficacy of the 6-week supplementation of *L. plantarum* PD01 in promoting the elimination of MPs and MP-derived chemicals in adults.

## Methods

### Trial design and oversight

This randomized, double-blind, placebo-controlled study was designed to evaluate probiotic-mediated clearance of MPs and MP-derived chemicals in overweight or obese participants based on prior evidence that microplastic accumulation drives obesity pathogenesis ^13,14^. This trial was registered with the U.S. Clinical Trials Registry (NCT07416214) and conducted in accordance with the Declaration of Helsinki. Study procedures were approved by the Ethics Committee of the School of Public Health, Fudan University (IRB#H2025029). Written informed consent was obtained from all participants prior to initiation of study.

### Participants

A total of 106 participants were recruited on November 2025. 96 participants were confirmed eligible based on the inclusion criteria and exclusion criteria. The inclusion criteria were as follow: (1) adults aged 18-65 years; (2) Body Mass Index (BMI) ≥ 24.0 kg/m^2^ or meeting the criteria for central obesity (waist circumference ≥ 90 cm for men, ≥ 85 cm for women); participants with mild abnormalities in blood lipids and liver function indicators will be prioritized. (3) permanent residents of the local area during the trial period, with no plans for long-term business trips or travel. (4) fully understand the research content and voluntarily sign the informed consent form. The exclusion criteria were as follows: (1) diagnosed congenital or acquired immunodeficiency diseases, severe allergic diseases, active gastrointestinal diseases, and other acute or chronic diseases requiring long-term treatment. (2) use of antibiotics, immunosuppressants, probiotics, prebiotics, symbiotic, or other drugs that clearly affect gut microbiota or gastrointestinal function within the 6 months prior to the trial. (3) regular intake of nutritional supplements (such as vitamins, fish oil, etc.) within the 6 months prior to the trial. (4) having bad lifestyle habits such as smoking or alcoholism. (5) women who are pregnant, lactating, or planning to become pregnant. (6) weight change exceeding 5% of body weight within the 3 months prior to the trial. (7) participation in or planning to participate in any other clinical intervention studies.

### Intervention

Eligible participants were randomly assigned in a 1:1 ratio to either the intervention group (n = 47) or the placebo group (n = 49) using a computer-generated randomization sequence created with SAS version 9.3. Randomization was managed by an administrator who was independent of this study and responsible for the distribution of the placebo or intervention. The randomization list, which assigned a unique randomization number to each intervention, was sealed in opaque envelope to ensure that participants, investigators, and data analysts were blinded to the intervention assignments throughout the study.

The intervention and placebo were prepared as solid beverage powders, identical in appearance, texture, and taste, and packaged in identical individual sachets. Each placebo sachet contained 2.0 g of fructooligosaccharides (FOS), each intervention contained 0.2 g of *L. plantarum* PD01 probiotic powder (1.0×10^10^ CFU) plus 1.8 g of FOS. During the 6-week intervention phase, participants were instructed to take one sachet with warm water (≤ 40 °C) after meals, three times daily.

Participants were asked to keep stable dietary habits and lifestyle to ensure consistent MPs exposure throughout the study. Dietary and lifestyle data were collected using questionnaire covering basic information, take-out and processed food consumption, drinking water records and lifestyle habits at baseline. The Food Frequency Questionnaire (FFQ) were collected at baseline, midpoint and endpoint. In addition, the 24-h dietary recalls for the past 3 day were conducted prior to assessments.

### Outcomes

The primary outcomes were changes of MP concentrations in feces and phthalate metabolite levels in urine. The concentration of MPs in human feces was determined using a Py-GC/MS method described previously with minor optimization to eliminate lipid interference ^12^. Fecal samples were lyophilized at -80 °C for 24 h and subsequently transferred into clean glass tubes. 10 mL of pure ethanol were added to the tube, and the samples were sonicated for 10 min. After centrifugation at 3000 rpm for 10 min, the supernatant was removed. This washing step was repeated twice with pure ethanol. Subsequently, the samples were washed with hexane using a similar procedure (centrifugation at 2000 rpm) and repeated twice to remove residual oil. The degreased samples were then digested with concentrated nitric acid at 60 °C for 24 h and subsequently concentrated to approximately 2 mL at 110-120 °C.

Concentrated samples were transferred into crucibles and dried for Py-GC/MS analysis.

At least 0.1 g of resulting samples were analyzed using a Frontier Lab EGA/PY3030D instrument (Shimadzu, Japan) equipped with an AS-2020E automatic sampler. Chromatographic separation was performed using an Ultra ALLOY+-5 capillary column, with MS data acquired in selected ion monitoring (SIM) mode. Raw data were processed using Qualitative Analysis 10.0 and MS Quantitative Analysis software, and polymers were identified against the F-Search MPs database. Ten target polymers were quantified using standard curves: polyamide 6 (PA6), polyamide 66 (PA66), polycarbonate (PC), polyethylene (PE), polyethylene terephthalate (PET), polymethyl methacrylate (PMMA), polypropylene (PP), polystyrene (PS), polyurethane (PU), and polyvinyl chloride (PVC). MP concentrations were normalized to the sample dry weight. Missing values (< 4%) were imputed via multiple imputation by chained equations with predictive mean matching (m = 5) using baseline demographics, followed by a recalculation of total polymer loads to ensure logical consistency. Concentrations below the limit of detection (LOD) were replaced with LOD/√2. The seven MPs detected in more than 50% of samples were included in the final statistical analysis. Quality control and analytical parameters were provided in Supplementary Appendix.

The urinary PAE metabolites were chromatographically separated on an Agilent Eclipse Plus C18 RRHD column (2.1 × 100 mm, 1.8 μm), and quantified using an SCIEX ExionLC AD ultra-high-performance liquid chromatography system coupled with an AB SCIEX 7500+ triple quadrupole mass spectrometer ^15^. Urinary creatinine concentrations were measured in the corresponding samples and used to adjust PAE metabolite levels. PAE metabolite concentrations below the limit of detection (LOD) were replaced with LOD/√2. Details for sample preparation and quality control were provided in Supplementary Appendix.

The secondary outcomes of body composition and biochemical parameters were assessed using standard methods ^16^. Body composition parameters included height, weight, BMI, body water, protein, minerals, body fat mass, soft-lean mass, fat-free mass, skeletal muscle mass, and body fat percentage. Biochemical parameters covered liver function, glucose metabolism, and lipid metabolism indicators, including triglycerides (TG) and high-density lipoprotein cholesterol (HDL-C). Detailed methods were provided in the Supplementary Appendix.

### Statistical analysis

Based on prior clinical data investigating the efficacy of *L. plantarum* in promoting pollutant excretion, an anticipated effect size (Cohen’s d) of 0.6 was used for sample size calculation ^17^. The significance level and power were set at 5% and 80%, respectively. A sample size of 44 evaluable participants was needed for each group. With an estimated dropout rate of 15 % during the trial period, a total of 104 participants was planned for recruitment.

Linear mixed-effects models (LMMs) were used to examine the effects of intervention on outcomes. In these models, time point (baseline vs. endpoint), treatment group (placebo vs. intervention), and their interaction (Group × Time) were included as fixed effects. To account for the within-subject correlation across repeated measurements, a subject-specific random intercept was included in the model. MPs and PAE metabolite levels were log_2_-transformed to normalize their distributions. Models estimating changes in MPs and PAE metabolites were adjusts for age, sex and time-varying BMI. For the longitudinal changes in body composition, models were adjusted for baseline age, sex, and time-varying dietary intake assessed via FFQs. For biochemical parameters, models were adjusted for age, sex, and time-varying BMI. The normality of model residuals was verified by QQ-plots. Within-group and between-group differences were evaluated using t-tests with Satterthwaite’s approximation for degrees of freedom. Statistical significance was defined as a two-sided *P* < 0.05. All statistical analyses and data visualizations were conducted in the R (version 4.4.3) using the lme4, lmerTest, and emmeans packages.

## Results

### Participants and characteristics

A total of 106 participants were recruited, 10 of whom were deemed ineligible based on the inclusion or exclusion criteria. The remaining participants were randomly allocated into either placebo or intervention group. Finally, 89 participants (44 in the placebo group and 45 in the intervention group) completed the study and included in the final analysis (Figure 1). The mean age of the participants in placebo group was 25.18 ± 4.55 years with 65.9% males, the mean age of the participants in intervention group was 24.66 ± 5.86 years with 66.7% males (Table 1). The body weight at baseline were 83.95 ± 12.69 and 84.02 ± 16.51 in placebo and intervention groups, respectively. The distribution of dietary MPs intake behaviors was equivalent between the two groups (Table 1).

**Table 1.**
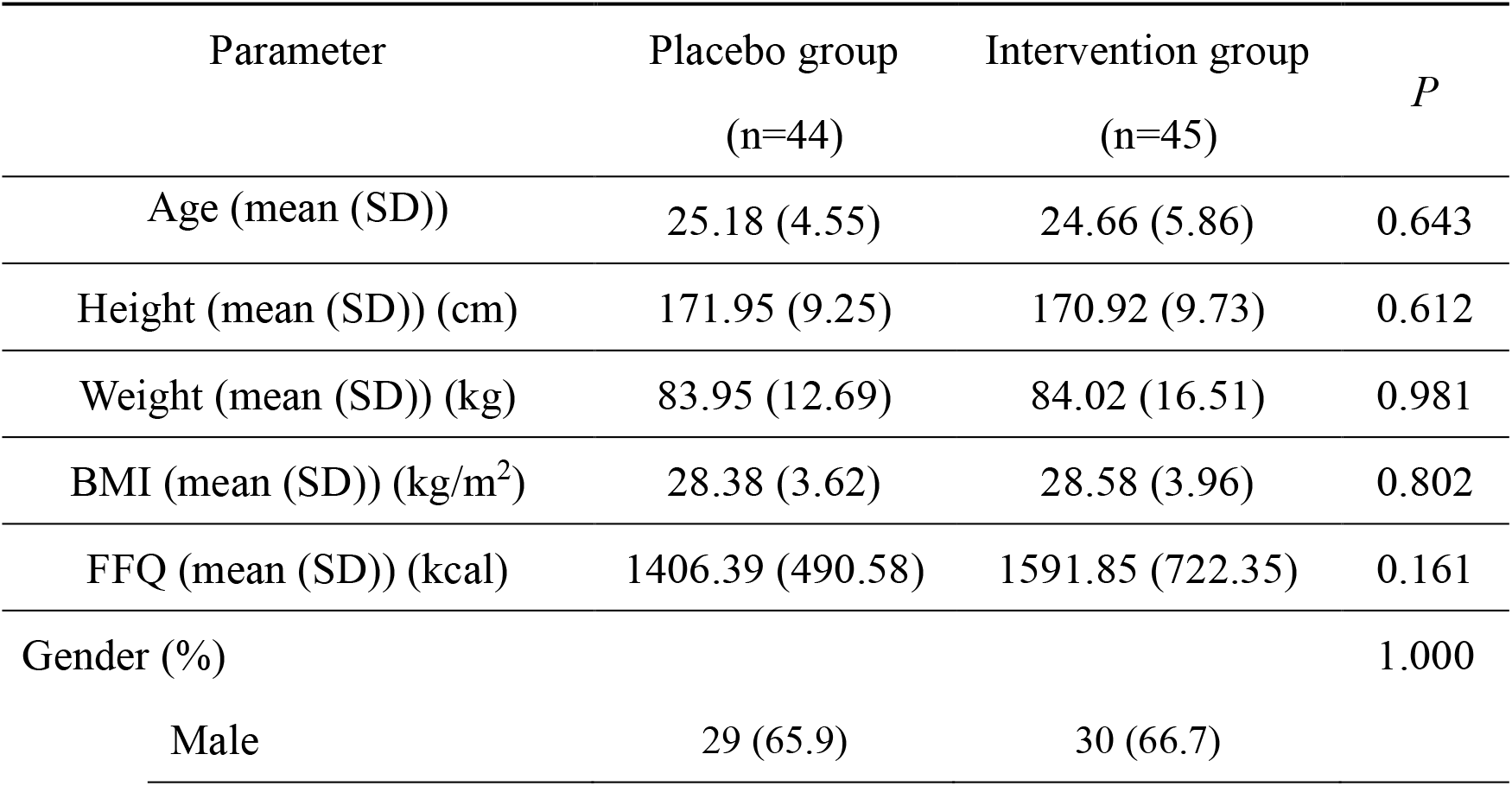
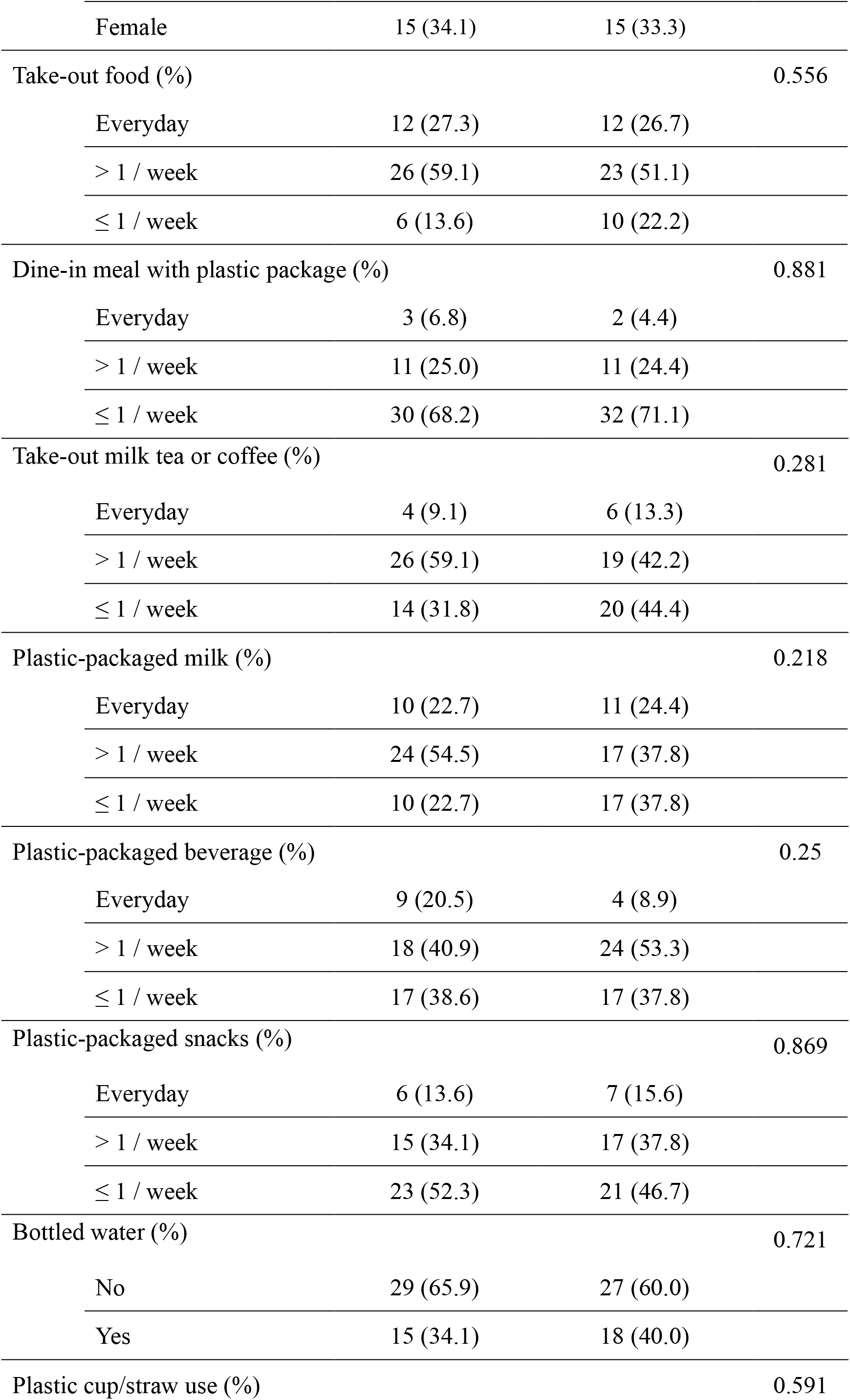
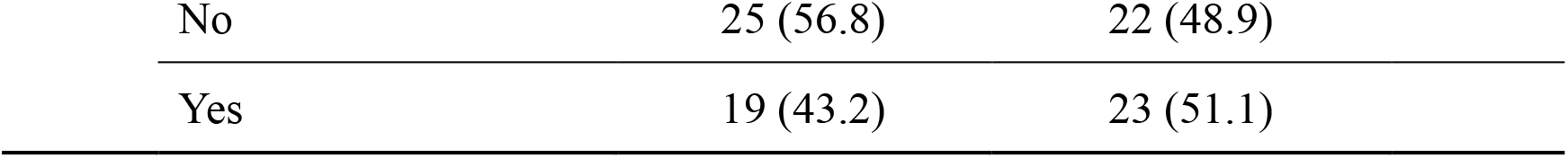
Baseline characteristics of the participants.

| Parameter | Placebo group<br>(n=44) | Intervention group<br>(n=45) | *P* |
| --- | --- | --- | --- |
| Age (mean (SD)) | 25.18 (4.55) | 24.66 (5.86) | 0.643 |
| Height (mean (SD)) (cm) | 171.95 (9.25) | 170.92 (9.73) | 0.612 |
| Weight (mean (SD)) (kg) | 83.95 (12.69) | 84.02 (16.51) | 0.981 |
| BMI (mean (SD)) (kg/m<sup>2</sup>) | 28.38 (3.62) | 28.58 (3.96) | 0.802 |
| FFQ (mean (SD)) (kcal) | 1406.39 (490.58) | 1591.85 (722.35) | 0.161 |
| Gender (%) |  |  | 1.000 |
| Male | 29 (65.9) | 30 (66.7) |  |
| Female | 15 (34.1) | 15 (33.3) |  |
| Take-out food (%) |  |  | 0.556 |
| Everyday | 12 (27.3) | 12 (26.7) |  |
| > 1 / week | 26 (59.1) | 23 (51.1) |  |
| ≤ 1 / week | 6 (13.6) | 10 (22.2) |  |
| Dine-in meal with plastic package (%) |  |  | 0.881 |
| Everyday | 3 (6.8) | 2 (4.4) |  |
| > 1 / week | 11 (25.0) | 11 (24.4) |  |
| ≤ 1 / week | 30 (68.2) | 32 (71.1) |  |
| Take-out milk tea or coffee (%) |  |  | 0.281 |
| Everyday | 4 (9.1) | 6 (13.3) |  |
| > 1 / week | 26 (59.1) | 19 (42.2) |  |
| ≤ 1 / week | 14 (31.8) | 20 (44.4) |  |
| Plastic-packaged milk (%) |  |  | 0.218 |
| Everyday | 10 (22.7) | 11 (24.4) |  |
| > 1 / week | 24 (54.5) | 17 (37.8) |  |
| ≤ 1 / week | 10 (22.7) | 17 (37.8) |  |
| Plastic-packaged beverage (%) |  |  | 0.25 |
| Everyday | 9 (20.5) | 4 (8.9) |  |
| > 1 / week | 18 (40.9) | 24 (53.3) |  |
| ≤ 1 / week | 17 (38.6) | 17 (37.8) |  |
| Plastic-packaged snacks (%) |  |  | 0.869 |
| Everyday | 6 (13.6) | 7 (15.6) |  |
| > 1 / week | 15 (34.1) | 17 (37.8) |  |
| ≤ 1 / week | 23 (52.3) | 21 (46.7) |  |
| Bottled water (%) |  |  | 0.721 |
| No | 29 (65.9) | 27 (60.0) |  |
| Yes | 15 (34.1) | 18 (40.0) |  |
| Plastic cup/straw use (%) |  |  | 0.591 |

|  |  |  |
| --- | --- | --- |
| No | 25 (56.8) | 22 (48.9) |
| Yes | 19 (43.2) | 23 (51.1) |

**Figure 1:**
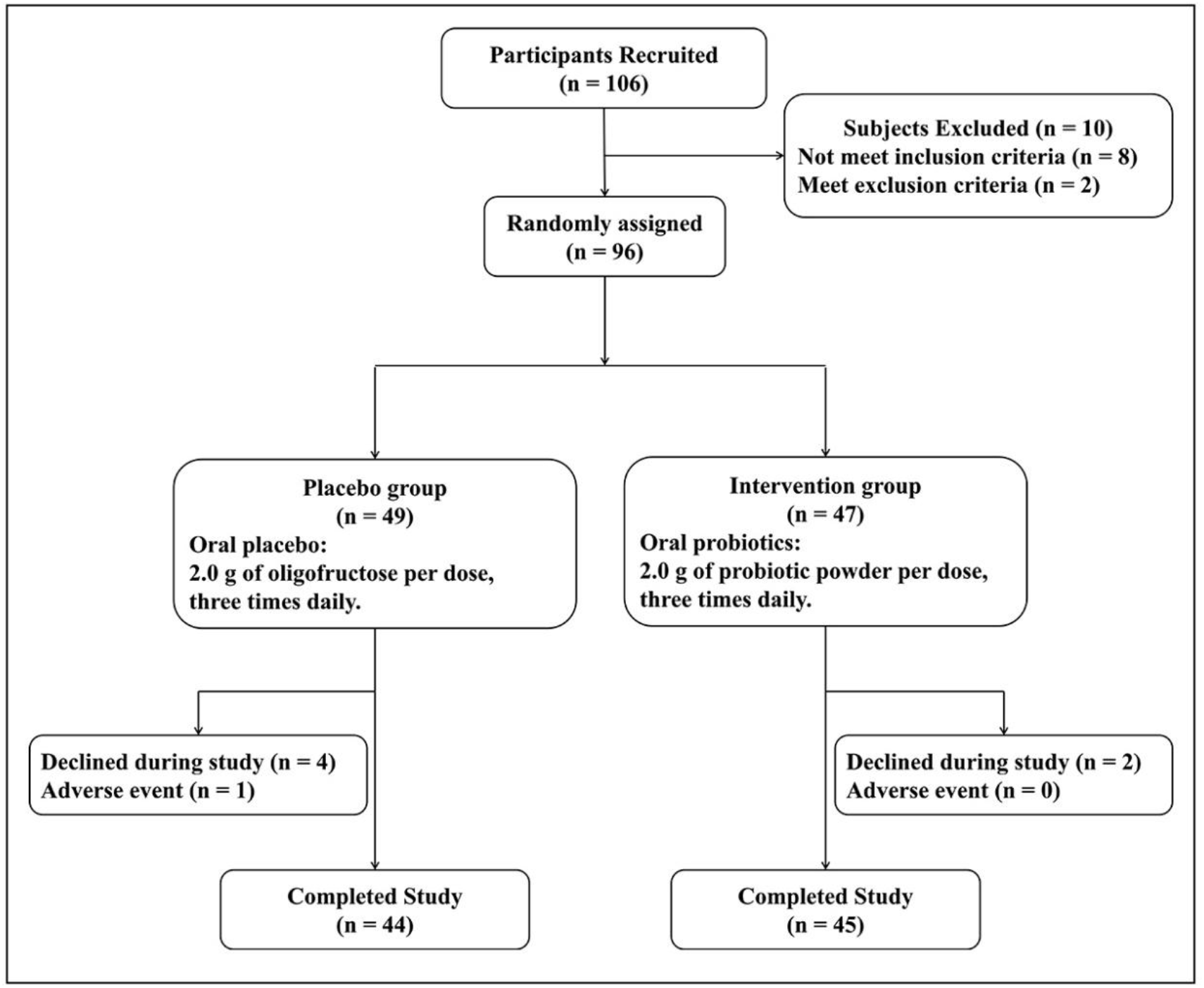
Flow diagram of RCT showing the process of random assignment, treatment administration, and the number of participants who completed the study.

During the study, only one adverse event was reported, occurring in a participant in the placebo group. This individual reported increased drowsiness and sleepiness, which led to discontinuation of the study treatment. The event resolved spontaneously and it was not related to treatment. No adverse events were reported in the intervention group.

### Intervention of *L. plantarum* PD01 increased fecal MPs excretion

Results from the LMMs are shown in Figure 2. After adjustment for age, sex and time-varying BMI, the fecal excretion of total MPs was significantly improved in the intervention group at both midpoint (estimated mean difference = 1.99; 95% CI: 0.78 to 3.21; interaction *P* = 0.001) and endpoint (1.25; 0.04 to 2.47; interaction *P* = 0.044), with significant Group × Time interactions (Table S1). There were significant interaction effects of intervention on PC amount at midpoint (3.19; 0.98 to 5.40; interaction *P* = 0.005) and endpoint (2.87; 0.66 to 5.08; interaction *P* = 0.011). A significant Group × Time interaction was also observed for PVC at midpoint (2.92; 0.60 to 5.23; interaction *P* = 0.014), though this differential trajectory was not sustained at endpoint (interaction *P* = 0.137). PE and PS increased significantly within the intervention group from baseline to midpoint (*P* = 0.005 and *P* = 0.002, respectively), and PE and PU increased significantly from baseline to endpoint (*P* = 0.042 and *P* = 0.044, respectively), but the Group × Time interactions did not reach significance (Table S1). No significant intervention effects were observed for PA66 and PP at either time point.

**Figure 2.**
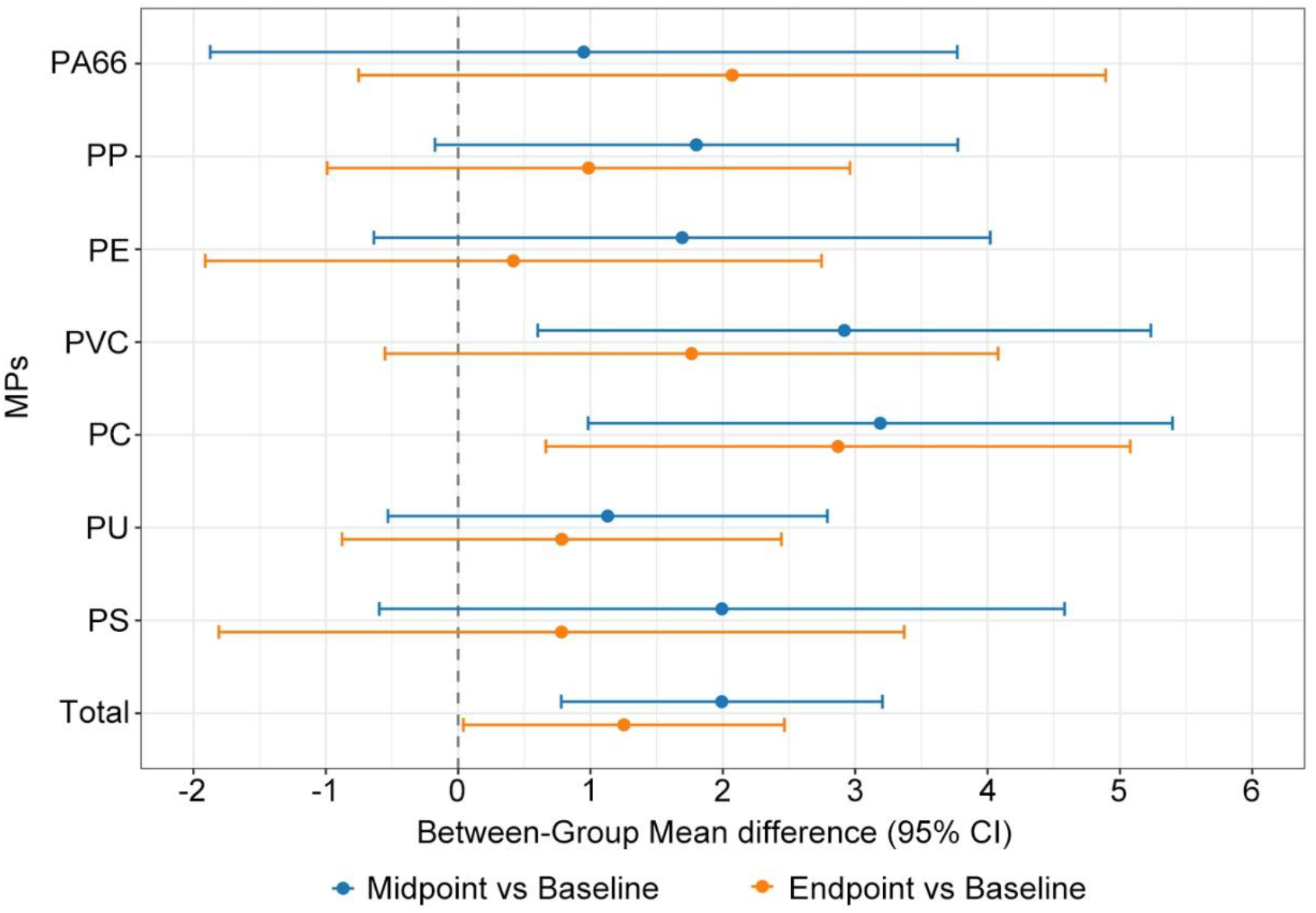
Changes in MPs after *L. plantarum* PD01 Intervention. Estimates are derived from linear mixed model that adjusts for age, sex and time-varying BMI. PC, polycarbonate. PVC, polyvinyl chloride. PA66, polyamide 66. PS, polystyrene. PP, polypropylene. PE, polyethylene. PU, polyurethane.

### Intervention of *L. plantarum* PD01 reduced urinary plasticizer levels

As shown in Figure 3, compared with placebo, the *L. plantarum* PD01 intervention significantly reduced MnBP (estimated mean difference = -0.96; 95% CI: -1.73 to -0.18; interaction *P* = 0.016), MEHHP (-0.91; -1.79 to -0.02; interaction *P* = 0.046), MECPP (-1.06; -2.09 to -0.03, interaction *P* = 0.045), and ΣDEHP (-0.97; -1.84 to -0.09, interaction *P* = 0.031) at endpoint (Table S1). Marginally significant interaction was observed for MMP (-0.65; -1.31 to 0.00; interaction *P* = 0.053) and MEOHP (-0.87; - 1.75 to 0.01; interaction *P* = 0.056).

**Figure 3.**
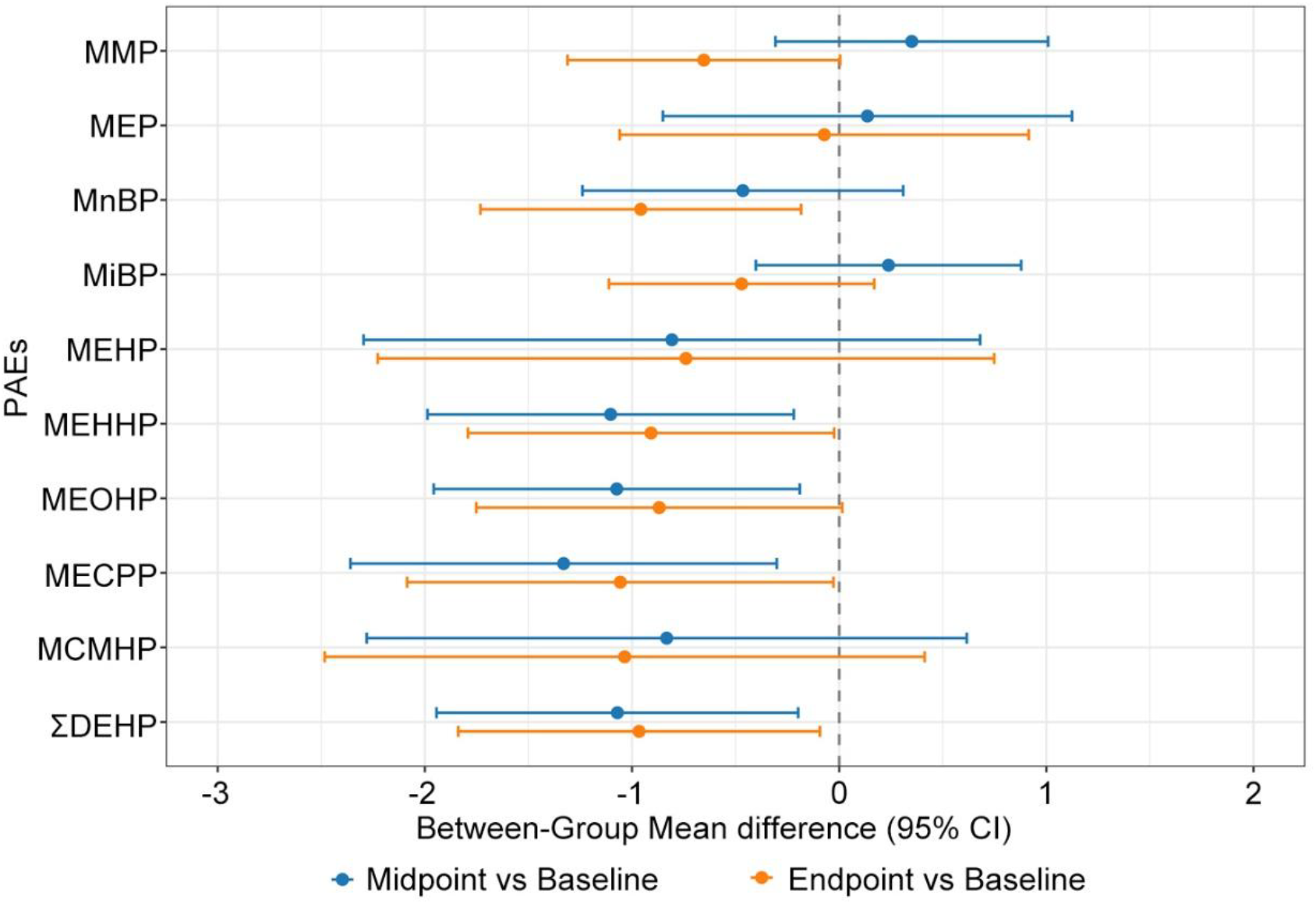
Changes in PAEs after *L. plantarum* PD01 Intervention. Estimates are derived from linear mixed model that adjusts for age, sex and time-varying BMI. MEP, mono-ethyl phthalate. MnBP, mono-n-butyl phthalate. MiBP, mono-isobutyl phthalate. MEHP mono(2-ethylhexyl) phthalate. MEHHP, mono(2-ethyl-5-hydroxyhexyl) phthalate. MEOHP, mono(2-ethyl-5-oxohexyl) phthalate. MECPP, mono(2-ethyl-5-carboxypentyl) phthalate. MCMHP, Mono[2-(carboxymethyl)hexyl] phthalate. ΣDEHP, total of di(2-ethylhexyl) phthalate (DEHP) metabolites.

### Effect of *L. plantarum* PD01 on body composition and biochemistry parameters

Based on LMMs adjusted with age, sex and time-varying dietary intake, the body weight changes were comparable between the intervention and placebo groups at both midpoint (Between-group mean difference = -0.27; 95% CI: -0.94 to -0.39; interaction *P* = 0.418) and endpoint (-0.22; -0.89 to 0.44; interaction *P* = 0.512) (Table S2). There were also no significant changes in other body composition parameters (Table S2). For the biochemistry outcomes, HDL-C levels were significantly higher in the intervention group at both time points (midpoint: 0.13; 95% CI: 0.04 to 0.22; interaction *P* = 0.006. endpoint: 0.18; 0.09 to 0.27; interaction *P* <0.001) (Table S3), and TC was significantly increased at endpoint (0.46; 0.06 to 0.85; interaction *P* = 0.025). No significant between-group differences were detected for other biochemical parameters.

## Discussion

Lactic acid bacteria promote pollutant elimination through physicochemical adsorption and restoration of the host intestinal barrier, thereby reducing systemic toxin retention ^18^. Here, we report the first randomized controlled trial demonstrating that *L. plantarum* PD01 strain effectively increases fecal MPs excretion and reduces the urinary levels of PAE metabolites in humans. These results are concordant with our findings from the murine model, in which *L. plantarum* PD01 prevented MPs translocation across the intestinal wall and achieved a 57.83% reduction in total MPs burden following dietary MPs mixture exposure ^12^. Unlike MPs, which can be excreted by feces, plasticizers released from MPs are readily absorbed from gastrointestinal tract and undergo hepatic metabolism ^19^. The observed reduction in urinary PAE metabolites following PD01 intervention reflects decreased entry of MPs and associated plasticizers into systemic circulation, consequently mitigating co-exposure to both pollutants and their associated synergistic toxicity ^7^

The prevalence and health risks of MPs and plasticizers have been broadly described in previous studies, yet few strategies have been proposed to counteract the plastic pollution pandemic, particularly in humans ^20^. A preliminary two-phase crossover study involving ten participants demonstrated that chitosan promoted fecal elimination by 45%, however, its conclusions were limited by the small sample size and the absence of a placebo-controlled arm ^21^. In the present study, we employed a more rigorous randomized controlled design with 96 participants and confirmed the promotion of total fecal MP excretion after 6-week PD01 intervention. The placebo FOS, a prebiotic known to stimulate gut motility and improve defecation ^22^, did not significantly enhance fecal MP excretion. This suggests that enhanced bowel movement alone is insufficient to eliminate MPs and that the MP-clearing effect of *L. plantarum* PD01 is specific. Collectively, these findings advance MP research beyond description and risk assessment toward prevention and therapeutic intervention.

Dietary and behavioral interventions that reduce plastic exposure can effectively decrease urinary PAEs metabolite concentrations ^23^. Significant reductions in exposure to phthalate metabolites were observed among mothers with young children following a one-month web-based behavioral intervention targeting dietary changes, personal care product use, and health behaviors ^24^. A seven-day low-plastic diet intervention combining plastic-free kitchenware, low-plastic personal-care products, and foods with minimal plastic contact reduced urinary plasticizer by 37.5% to 59.7% ^8^. In our study, we achieved reduction in urinary PAEs metabolite levels by promoting MPs excretion rather than through plastic-free food or behavioral interventions. Previous preclinical studies indicate that supplementation of PD01 may further enhance the PAEs clearance by direct adsorption of phthalates ^25^. Given that dietary and behavioral strategies are difficult to implement uniformly across diverse populations, this study provides a more feasible and low-cost approach for clearing plastic pollutants in humans.

The extent to which ingested microplastics are retained, translocated, or excreted in the gastrointestinal tract depends on polymer composition, particle size and surface properties ^26^. Consistent with this heterogeneity, the response to *L. plantarum* PD01 varied across polymer types in our trial. PC showed the largest increase in fecal excretion, with a significant Group × Time interaction. Other polymers, although not reaching significant interaction effects individually, exhibited directionally consistent increases over time within the intervention group and relative to placebo. The accumulation of these polymer-specific effects yielded a significant Group × Time interaction for total MPs. Thus, despite variability in single-polymer responses, *L. plantarum* PD01 appears to exert a broad-spectrum effect on fecal MP excretion. Whether this efficacy extends to populations with different MP exposure warrants further investigation in more diverse cohorts.

Previous animal studies have proposed MPs as environmental obesogens that disrupt gut-liver-adipose axis and metabolic homeostasis ^13,14^. However, although our intervention effectively enhanced MP and phthalate elimination, it did not significantly alter body weight or body composition. This discrepancy may reflect differences between animals and humans in the contribution of MP accumulation to obesity development; alternatively, a longer intervention period that sustains low MPs burden may be required to repair MP-induced metabolic disruption. As substantial evidence on MP toxicity still derives from animal models, extrapolation of such findings to human health risks requires a more cautious interpretation.

In summary, compared with the placebo group, the 6-week supplementation with *L. plantarum* PD01 effectively promoted the elimination of MPs and PAEs from the body. Our findings provide the first clinical evidence that plastic-associated pollutant burden can be lowered without dietary or behavioral restrictions, bridging the critical animal-to-human translation gap in human toxicology and risk assessment.

## Author Approvals

All authors have seen and approved the manuscript, and that it hasn’t been accepted or published elsewhere.

## Data Availability Statement

The data presented in this study are available on request from the corresponding author.

## Notes

### Competing Interest Statement

The authors have declared no competing interest.

### Clinical Trial

NCT07416214

### Author Declarations

This randomized, double-blind, placebo-controlled study was registered with the U.S. Clinical Trials Registry (NCT07416214) and conducted in accordance with the Declaration of Helsinki. Study procedures were approved by the Ethics Committee of the School of Public Health, Fudan University (IRB#H2025029). Written informed consent was obtained from all participants prior to initiation of study.

### Summary of Updates

Results of fecal MPs were updated. Results of PAEs were updated.

## References

1. Akhtar A, Xu H, Gulzar S, et al. Lifetime exposure to microplastics: From consumption to distribution in the body. Journal of Hazardous Materials Advances 2026;21:101077. DOI: 10.1016/j.hazadv.2026.101077.

2. Zhu L, Kang Y, Ma M, et al. Tissue accumulation of microplastics and potential health risks in human. Science of The Total Environment 2024;915:170004. DOI: 10.1016/j.scitotenv.2024.170004.

3. Kumari M, Pulimi M. Phthalate esters: occurrence, toxicity, bioremediation, and advanced oxidation processes. Water Science and Technology 2023;87(9):2090–2115. DOI: 10.2166/wst.2023.119.

4. Chang W-H, Herianto S, Lee C-C, Hung H, Chen H-L. The effects of phthalate ester exposure on human health: A review. Science of The Total Environment 2021;786:147371. DOI: 10.1016/j.scitotenv.2021.147371.

5. Liu F-f, Liu G-z, Zhu Z-l, Wang S-c, Zhao F-f. Interactions between microplastics and phthalate esters as affected by microplastics characteristics and solution chemistry. Chemosphere 2019;214:688–694. DOI: 10.1016/j.chemosphere.2018.09.174.

6. Zhang F, Chen H, Liu Y, Wang M. Phthalate acid ester release from microplastics in water environment and their comparison between single and competitive adsorption. Environmental Science and Pollution Research 2023;30(56):118964–118975. DOI: 10.1007/s11356-023-30720-4.

7. Liu H, Zheng D, Liu X, Hou J, Wu Q, Li Y. Environmental microplastic and phthalate esters co-contamination, interrelationships, co-toxicity and mechanisms. A review. Environmental Geochemistry and Health 2024;46(12):525. DOI: 10.1007/s10653-024-02309-4.

8. Harray AJ, Lucas AD, Herrmann SE, et al. Low-plastic diet and urinary levels of plastic-associated phthalates and bisphenols: the randomized controlled PERTH Trial. Nature Medicine 2026;32(5):1871–1883. DOI: 10.1038/s41591-026-04324-7.

9. Demarquoy J. Microplastics and probiotics: Mechanisms of interaction and their consequences for health. AIMS Microbiology 2025;11(2):388–409. DOI: 10.3934/microbiol.2025018.

10. Xiao R, Yu Y, Ning D, et al. A detoxifying multi-strain probiotic formula attenuates toxicity induced by heavy metals and phthalates. Food Research International 2026;233:119002. DOI: 10.1016/j.foodres.2026.119002.

11. Shi L, Wu C, Wang Y, et al. Lactobacillus plantarum reduces polystyrene microplastic induced toxicity via multiple pathways: A potentially effective and safe dietary strategy to counteract microplastic harm. Journal of Hazardous Materials 2025;489:137669. DOI: 10.1016/j.jhazmat.2025.137669.

12. Zhao L, Feng Y, Li J, et al. Lactiplantibacillus plantarum PD01 from high excretion donor functions as a natural defense against microplastic toxicity. npj Science of Food 2026. DOI: 10.1038/s41538-026-01045-6.

13. Zhao J, Adiele N, Gomes D, et al. Obesogenic polystyrene microplastic exposures disrupt the gut-liver-adipose axis. Toxicol Sci 2024;198(2):210–220. (In eng). DOI: 10.1093/toxsci/kfae013.

14. Huang H, Wei F, Qiu S, Xing B, Hou J. Polystyrene microplastics trigger adiposity in mice by remodeling gut microbiota and boosting fatty acid synthesis. Sci Total Environ 2023;890:164297. (In eng). DOI: 10.1016/j.scitotenv.2023.164297.

15. Chen M, Tao L, Collins EM, Austin C, Lu C. Simultaneous determination of multiple phthalate metabolites and bisphenol-A in human urine by liquid chromatography–tandem mass spectrometry. Journal of Chromatography B 2012;904:73–80. DOI: 10.1016/j.jchromb.2012.07.022.

16. Haganes KL, Silva CP, Eyjólfsdóttir SK, et al. Time-restricted eating and exercise training improve HbA1c and body composition in women with overweight/obesity: A randomized controlled trial. Cell Metabolism 2022;34(10):1457-1471.e4. DOI: 10.1016/j.cmet.2022.09.003.

17. Ji W, Saulnier DM, Zhang L, et al. Effects of Lactiplantibacillus plantarum DSM 33464 in children with elevated blood lead levels: a randomized, double-blind, placebo-controlled study. Frontiers in Nutrition 2025;Volume 12 - 2025 (Clinical Trial) (In English). DOI: 10.3389/fnut.2025.1641839.

18. Shi L, Wu C, Wang Y, et al. Lactobacillus plantarum reduces polystyrene microplastic induced toxicity via multiple pathways: A potentially effective and safe dietary strategy to counteract microplastic harm. J Hazard Mater 2025;489:137669. (In eng). DOI: 10.1016/j.jhazmat.2025.137669.

19. Kluwe WM. Overview of phthalate ester pharmacokinetics in mammalian species. Environ Health Perspect 1982;45:3–9. (In eng). DOI: 10.1289/ehp.82453.

20. Donisi I, Colloca A, Anastasio C, Balestrieri ML, D’Onofrio N. Micro(nano)plastics: an Emerging Burden for Human Health. Int J Biol Sci 2024;20(14):5779–5792. (In eng). DOI: 10.7150/ijbs.99556.

21. Casella C, Cornelli U, Ballaz S, Recchia M, Zanoni G, Ramos-Guerrero L. Preliminary Study on PCC-Chitosan’s Ability to Enhance Microplastic Excretion in Human Stools from Healthy Volunteers. Foods 2025;14(13) (In eng). DOI: 10.3390/foods14132190.

22. Yen CH, Kuo YW, Tseng YH, Lee MC, Chen HL. Beneficial effects of fructo-oligosaccharides supplementation on fecal bifidobacteria and index of peroxidation status in constipated nursing-home residents--a placebo-controlled, diet-controlled trial. Nutrition 2011;27(3):323–8. (In eng). DOI: 10.1016/j.nut.2010.02.009.

23. Sieck NE, Bruening M, van Woerden I, Whisner C, Payne-Sturges DC. Effects of Behavioral, Clinical, and Policy Interventions in Reducing Human Exposure to Bisphenols and Phthalates: A Scoping Review. Environ Health Perspect 2024;132(3):36001. (In eng). DOI: 10.1289/ehp11760.

24. Kim JH, Kwak JM, Kang H. Web-based behavioral intervention to reduce exposure to phthalate metabolites, bisphenol A, triclosan, and parabens in mothers with young children: A randomized controlled trial. International Journal of Hygiene and Environmental Health 2021;236:113798. DOI: 10.1016/j.ijheh.2021.113798.

25. Goyal SP, Saravanan C. An insight into the critical role of gut microbiota in triggering the phthalate-induced toxicity and its mitigation using probiotics. Science of The Total Environment 2023;904:166889. DOI: 10.1016/j.scitotenv.2023.166889.

26. Dávalos A, Martín-Santiago V, López de Las Hazas Mc. Microplastics and Nanoplastics in the Human Intestine: A Review of Health Implications and Possible Dietary Interventions. Adv Nutr 2026;17(5):100624. (In eng). DOI: 10.1016/j.advnut.2026.100624.

